# Acute kidney injury is associated with severe and fatal outcomes in patients with Coronavirus disease 2019 (COVID-19) infection: a systematic review and meta-analysis of observational studies

**DOI:** 10.1101/2020.08.27.20183632

**Authors:** Mohammad Parohan, Sajad Yaghoubi, Mahmoud Djalali, Asal Seraji, Mohammad Hassan Javanbakht, Zahra Mousavi

## Abstract

Coronavirus disease 2019 (COVID-19) is a pandemic impacting 213 countries and territories with more than 17,918,582 cases worldwide. Kidney dysfunction has been reported to occur in severe and death cases. This meta-analysis was done to summarize available studies on the association between acute kidney injury and severity of COVID-19 infection. Online databases including Web of Science, PubMed/Medline, Cochrane Library, Scopus and Google Scholar were searched to detect relevant articles up to 1 July 2020, using relevant keywords. To pool data, a random- or fixed-effects model was used based on the heterogeneity between studies. In total, 50 studies with 8,180 COVID-19 confirmed cases (severe cases=1,823 and death cases=775), were included in this meta-analysis. Higher serum levels of creatinine (weighted mean difference (WMD) for disease severity=5.47 μmol/L, 95% CI=2.89 to 8.05, P<0.001 and WMD for mortality=18.32 μmol/L, 95% CI=12.88 to 23.75, P<0.001), blood urea nitrogen (BUN) (WMD for disease severity=1.10 mmol/L, 95% CI=0.67 to 1.54, P<0.001 and WMD for mortality=3.56 mmol/L, 95% CI=2.65 to 4.48, P<0.001) and lower levels of estimated glomerular filtration rate (eGFR) (WMD for disease severity=-15.34 mL/min/1.73 m^2^, 95% CI=-18.46 to -12.22, P<0.001 and WMD for mortality=-22.74 mL/min/1.73 m^2^, 95% CI=-27.18 to -18.31, P<0.001) were associated with a significant increase in the severity and mortality of COVID-19 infection. Acute kidney injury, as assessed by kidney biomarkers (serum creatinine, BUN and eGFR), was associated with severe outcome and death from COVID-19 infection.

## Introduction

Coronavirus disease 2019 (COVID-19) is a global pandemic impacting 213 countries, areas and territories with a total of 17,918,582 confirmed cases and 686,703 deaths worldwide ^1^. As the COVID-19 outbreak expands, identification of clinical and laboratory predictors for severe and fatal outcomes are essential to enable risk stratification, guide public health recommendations and optimize allocation of limited hospital resources in the ongoing pandemic.

Genome sequencing showed that the COVID-19 belongs to β genus of Coronavirus, which also includes Middle East respiratory syndrome (MERS) and severe acute respiratory syndrome (SARS) ^2^. Similar to SARS, COVID-19 infection is caused by binding of the virus to the angiotensin-converting enzyme 2 (ACE2) receptors ^3^. ACE2 receptors are highly expressed in the lungs, kidneys, heart, vascular endothelium, liver and intestinal epithelium, providing a mechanism for the multi-organ dysfunction that can be seen with COVID-19 infection ^4, 5^.

Multiple observational studies have reported the clinical features and laboratory findings associated with acute kidney injury in patients with COVID-19 infection ^6-55^. In this study, the laboratory findings and mechanism of kidney dysfunction caused by COVID-19 were summarized.

## Methods

### Study protocol

A systematic literature search was conducted according to the Preferred Reporting Items for Systematic Reviews and Meta-Analyses (PRISMA) guidelines ^56^.

### Search strategy

We conducted a systematic search using the online databases of Web of Science, PubMed/Medline, Cochrane Library, Scopus and Google Scholar for relevant articles up to 1 July 2020. The following medical subject headings (MeSH) and non-MeSH terms were used: (“Novel Coronavirus” OR “2019‐nCoV” OR “Coronavirus disease 2019” OR “COVID-19” OR “Severe acute respiratory syndrome Coronavirus 2” OR “SARS-CoV-2”) AND (“estimated Glomerular filtration rate” OR “eGFR” OR “Creatinine clearance” OR “Blood Urea Nitrogen” OR “BUN” OR “Creatinine” OR “Kidney diseases” OR “Acute Kidney injury”). The systematic search was done by two reviewers (MP and SY). The reference lists of the relevant articles were also searched to identify missed studies. No restriction was applied to publication year and language. The search results were downloaded into an EndNote library (version X8, Thomson Reuters, Philadelphia, USA) and titles/abstracts assessed for eligibility. The keywords and search strategies are presented in Supplementary Table 1.

### Eligibility Criteria

Studies were included in this systematic review and meta-analysis if they met the following inclusion criteria: (1) observational studies (either retrospective or prospective); (2) studies assessing the association between kidney biomarkers (including eGFR, BUN and serum creatinine) and severe outcome or death from COVID-19 infection and reported mean (SD) or median (IQR) for these biomarkers. Reviews, expert opinion articles, books and theses were excluded.

### Data extraction and assessment for study quality

Two reviewers (MP and AS) extracted the following data from the included articles: publication year, author’s name, study design, demographic characteristics including age and gender, sample size, outcome assessment methods and kidney biomarkers including eGFR, BUN and creatinine.

The Newcastle-Ottawa Scale was used for assessing the quality and risk of bias of included articles ^57^. Based on this criteria, a maximum of nine points can be awarded to each article. In this meta-analysis, articles with the Newcastle–Ottawa Scale score of ≥ 5 were considered as high quality studies.

### Statistical analysis

Mean (SD) or median (IQR) for eGFR, BUN and serum creatinine were used to estimate the effect size. The fixed or random-effect model was performed depending on the heterogeneity between studies. Heterogeneity between studies was assessed using the Cochrane Q test and I^2^ statistics ^58^. The publication bias was assessed by the Egger’s regression test ^59^.The sensitivity analysis was used to evaluate the effect of each study on the pooled effect size. Statistical analyses were performed using the Stata 14 software package (Stata Corp, College Station, TX, USA).

## Results

### Search results

Overall, 623 articles were identified in our systematic literature search. Of these, 41 non-English, 215 duplicates, 83 reviews and 223 articles that did not fulfill our inclusion criteria were excluded, leaving 61 publications for further evaluation. Out of remaining 61 papers, 11 were excluded because of the following reason: did not report mean (SD) or median (IQR) for kidney biomarkers. Finally, we included 50 articles in this meta-analysis (Figure 1).

**Fig. 1.**
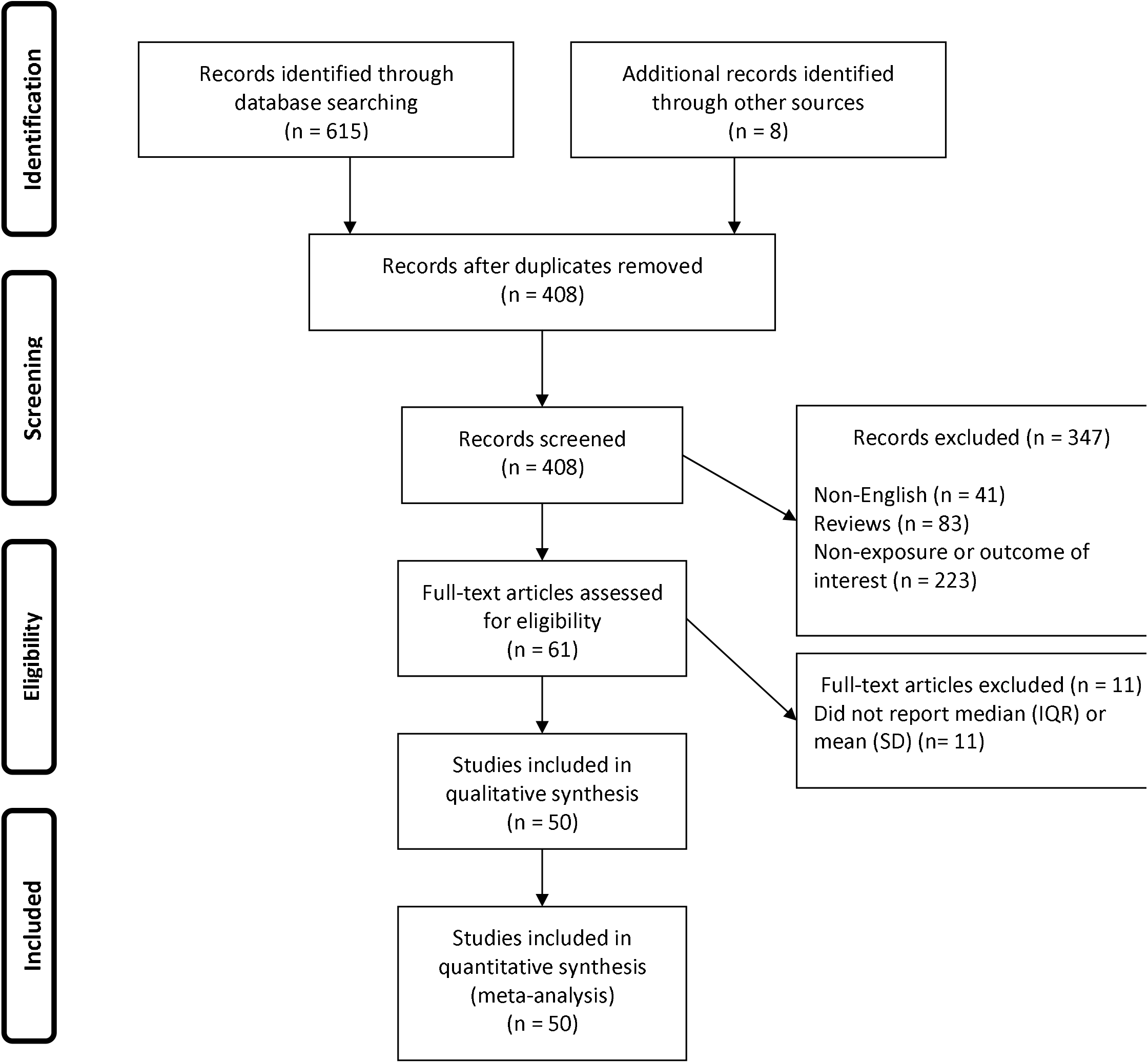
Flow chart of study selection

### Study characteristics

Forty-six studies were conducted in China ^6-17, 19, 20, 22-28, 30-34, 36-43, 45-55, 60^, one in United States ^29^, one in United Kingdom^44^, one in Japan^35^ and one in Italy^21^. Forty-seven studies used retrospective design ^6, 7, 9-15, 17, 19, 20, 22-55, 60^ and three studies used prospective design ^8, 16, 21^. The mean age in severe and non-severe patients were 64.4 and 52.1 years, respectively. The sample size of studies ranged from 10 to 671 participants. All studies used real-time reverse transcriptase–polymerase chain reaction (RT-PCR) method to confirm COVID-19 infection. The characteristics of the studies are presented in Table 1.

**Table 1.**
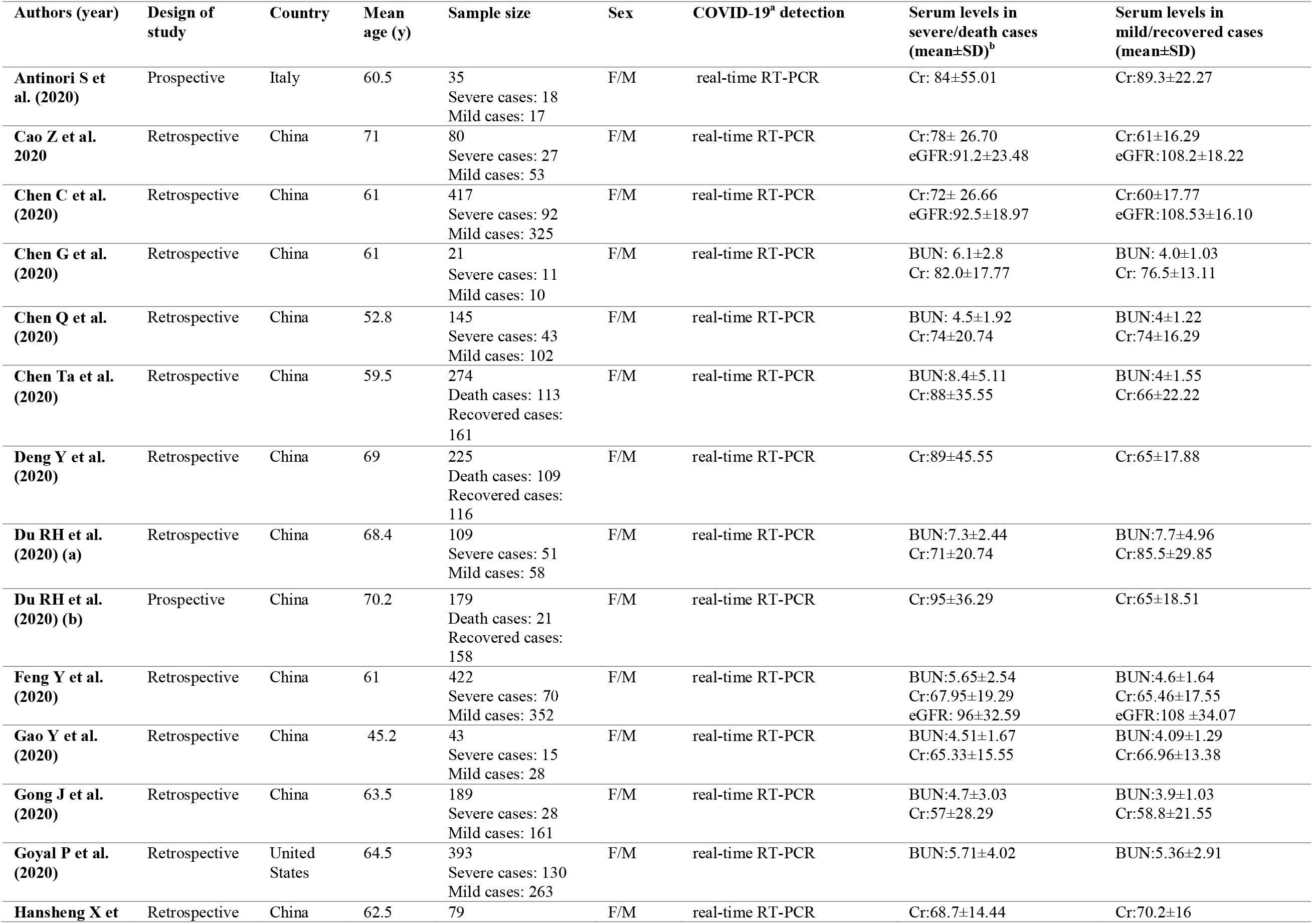
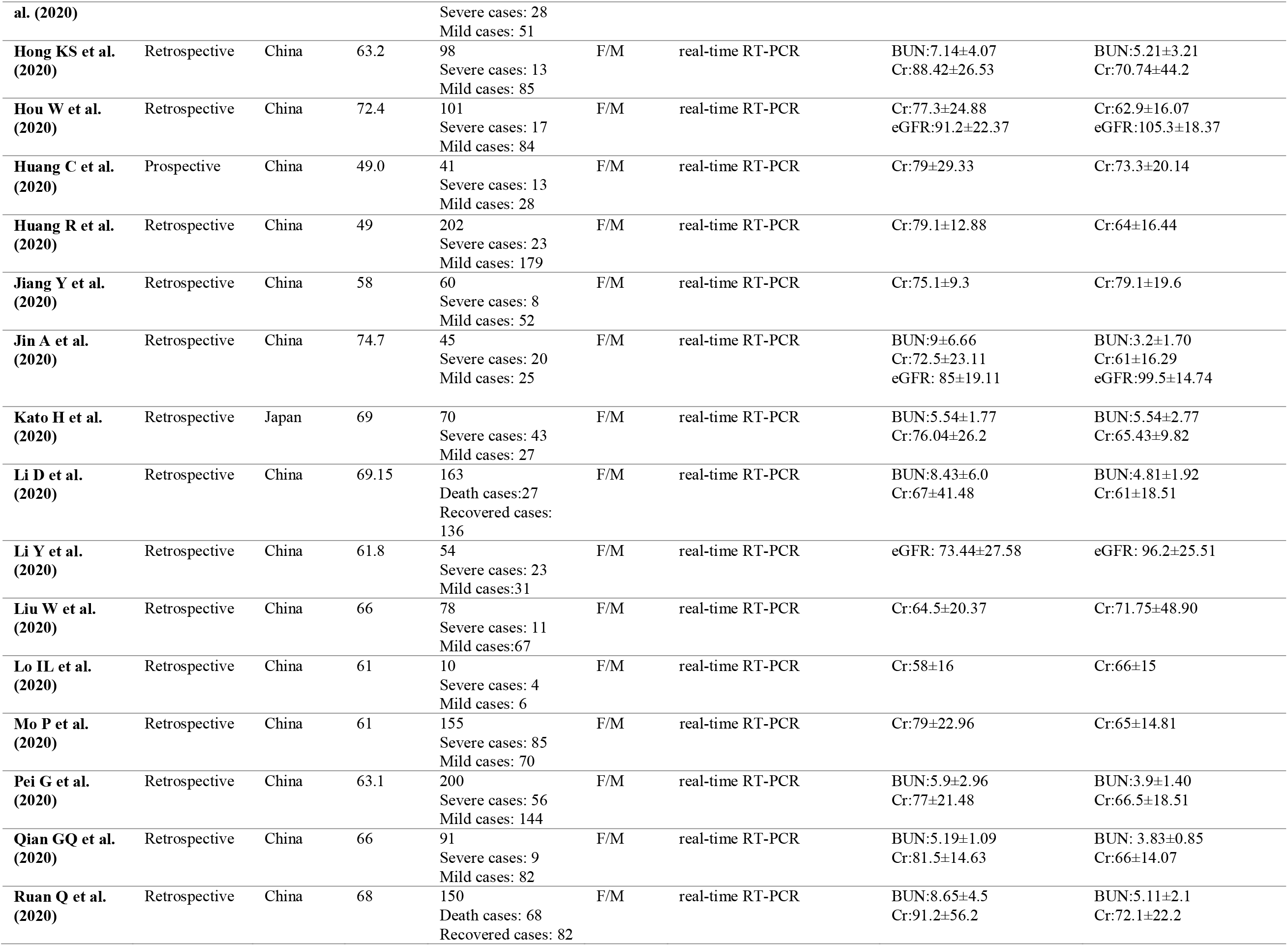
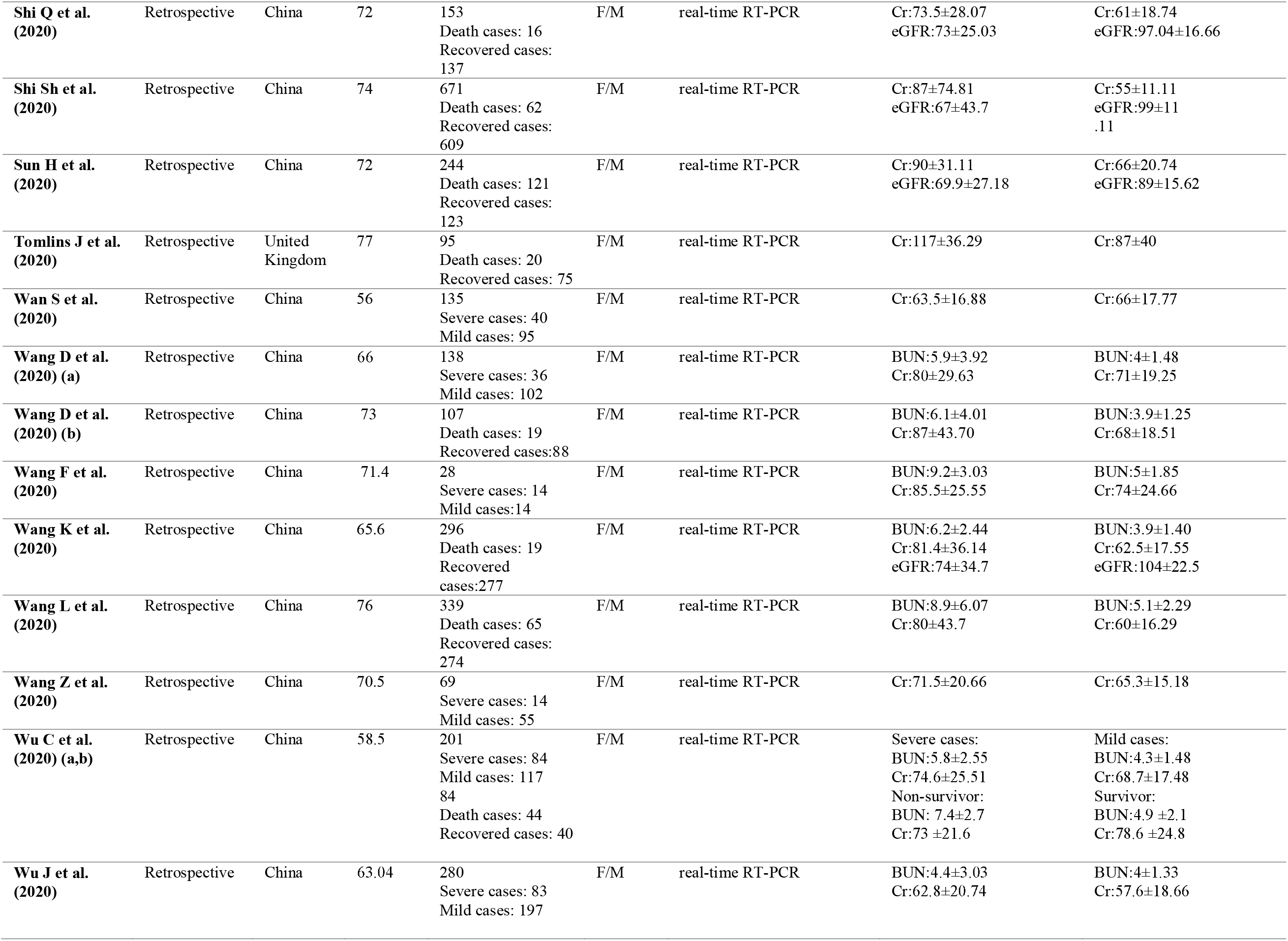
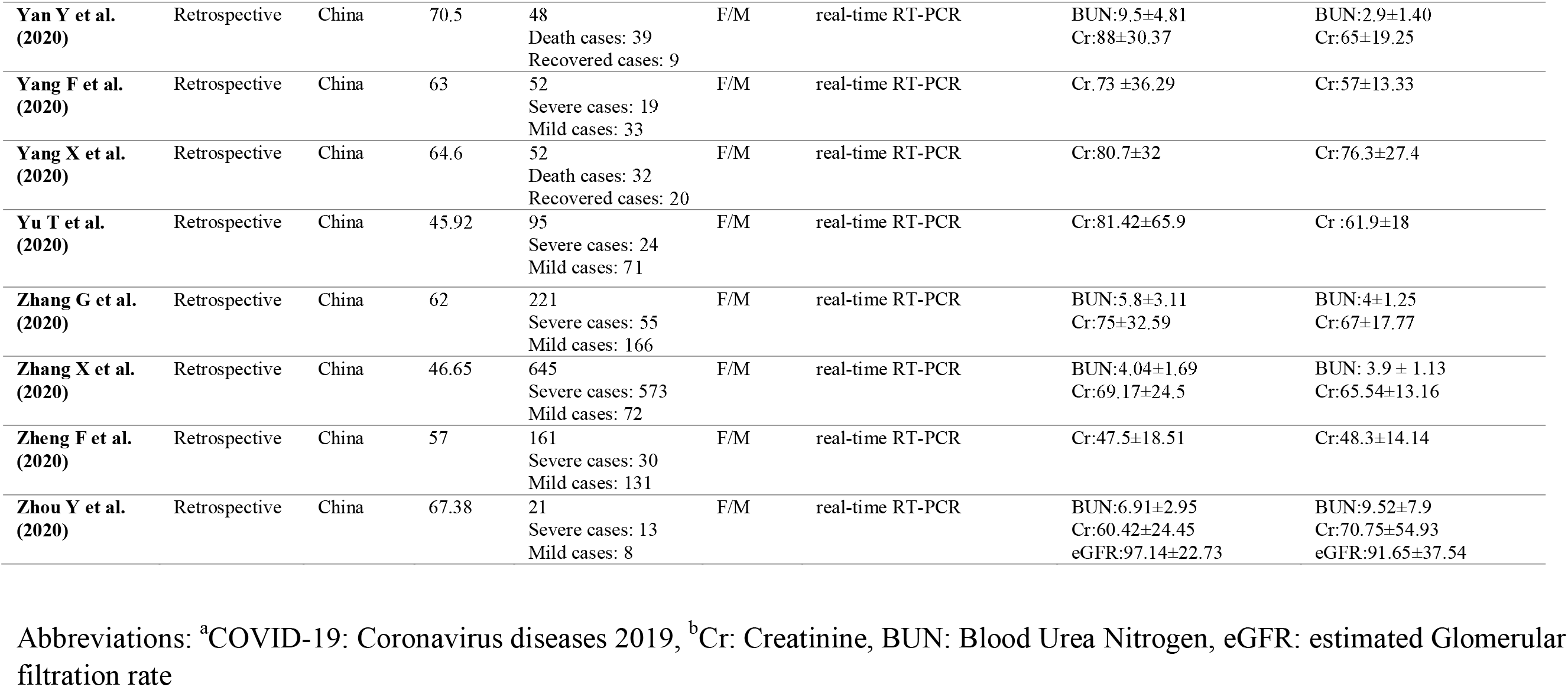
Characteristics of studies included in the meta-analysis.

### Serum levels of creatinine, BUN, eGFR and severity of COVID-19 infection

In the pooled estimate of 36 studies ^6, 8-15, 17, 19, 21-24, 26-35, 37-40, 46, 49, 51, 53-55, 60^ with 5,104 COVID-19 confirmed patients (severe cases = 1,796 and non-severe cases = 3,308), it was shown that higher serum levels of creatinine (weighted mean difference = 5.47 μmol/L, 95% CI = 2.89 to 8.05, P<0.001, I^2^ = 64.5%, P_heterogeneity_ <0.001) (Figure 2) and BUN (weighted mean difference = 1.10 mmol/L, 95% CI = 0.67 to 1.54, P<0.001, I^2^ = 77.3%, P_heterogeneity_ <0.001) (Figure 3), were associated with a significant increase in the severity of COVID-19 infection. In addition, combined results from the fixed-effects model showed that lower levels of eGFR (weighted mean difference = -15.34 mL/min/1.73 m^2^, 95% CI = -18.46 to -12.22, P<0.001, I^2^ = 0.0%, P_heterogeneity_ = 0.689) (Figure 4), significantly increased severity of the disease.

**Fig. 2.**
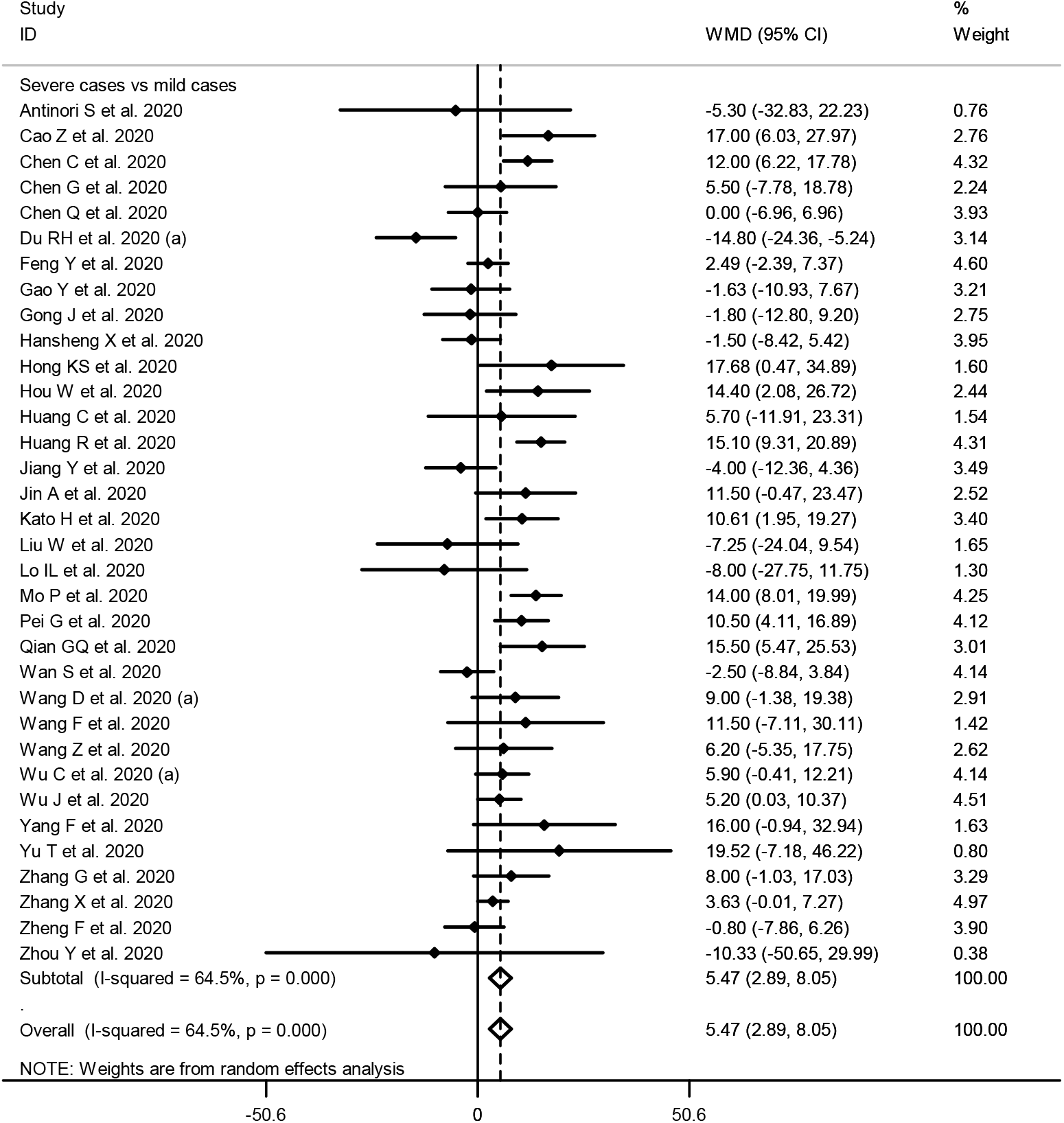
Forest plot for the association between serum levels of creatinine and severe outcome from COVID-19 infection using random-effects model

**Fig. 3.**
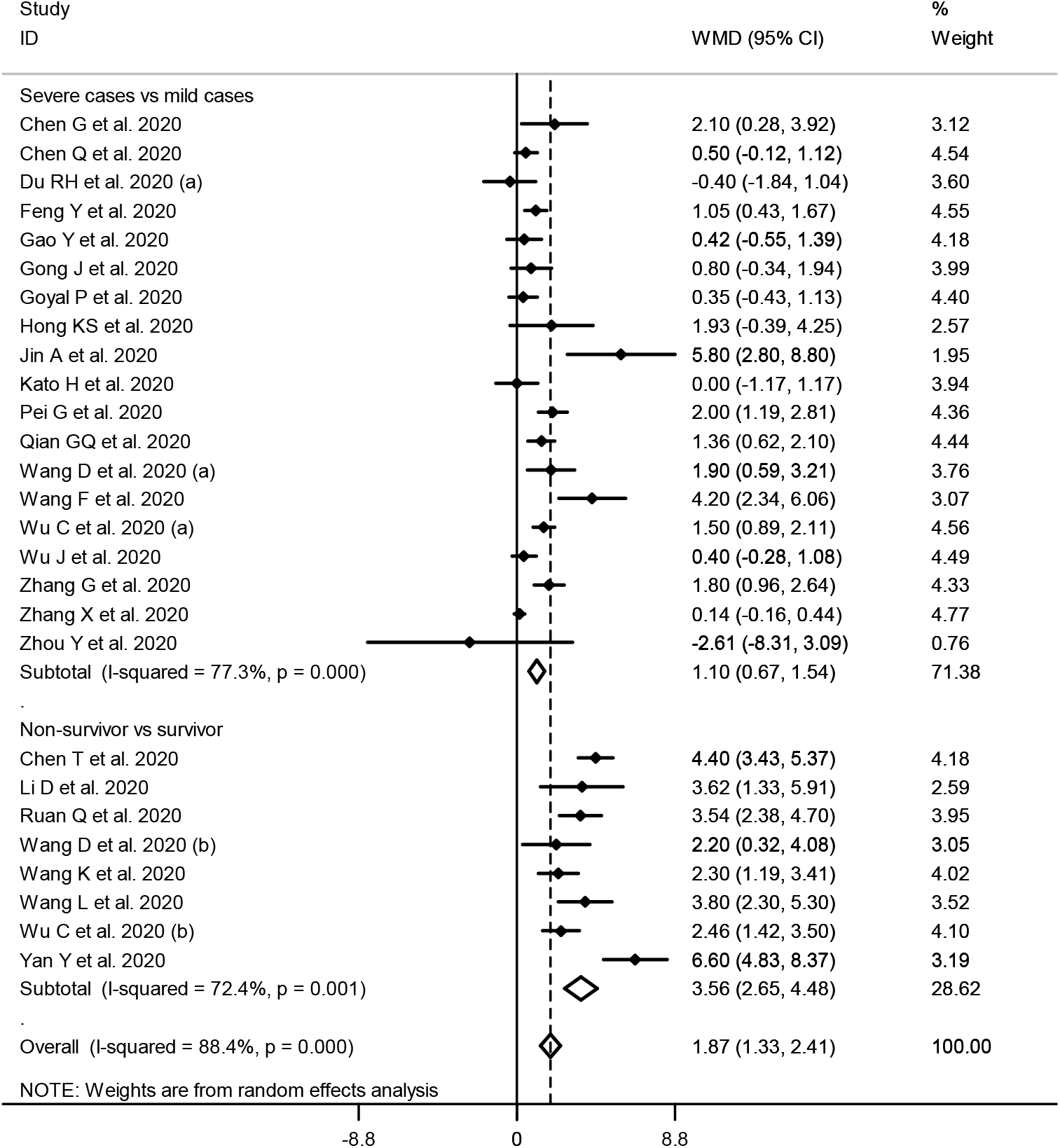
Forest plot for the association between BUN levels and severe outcome or death from COVID-19 infection using random-effects model

**Fig. 4.**
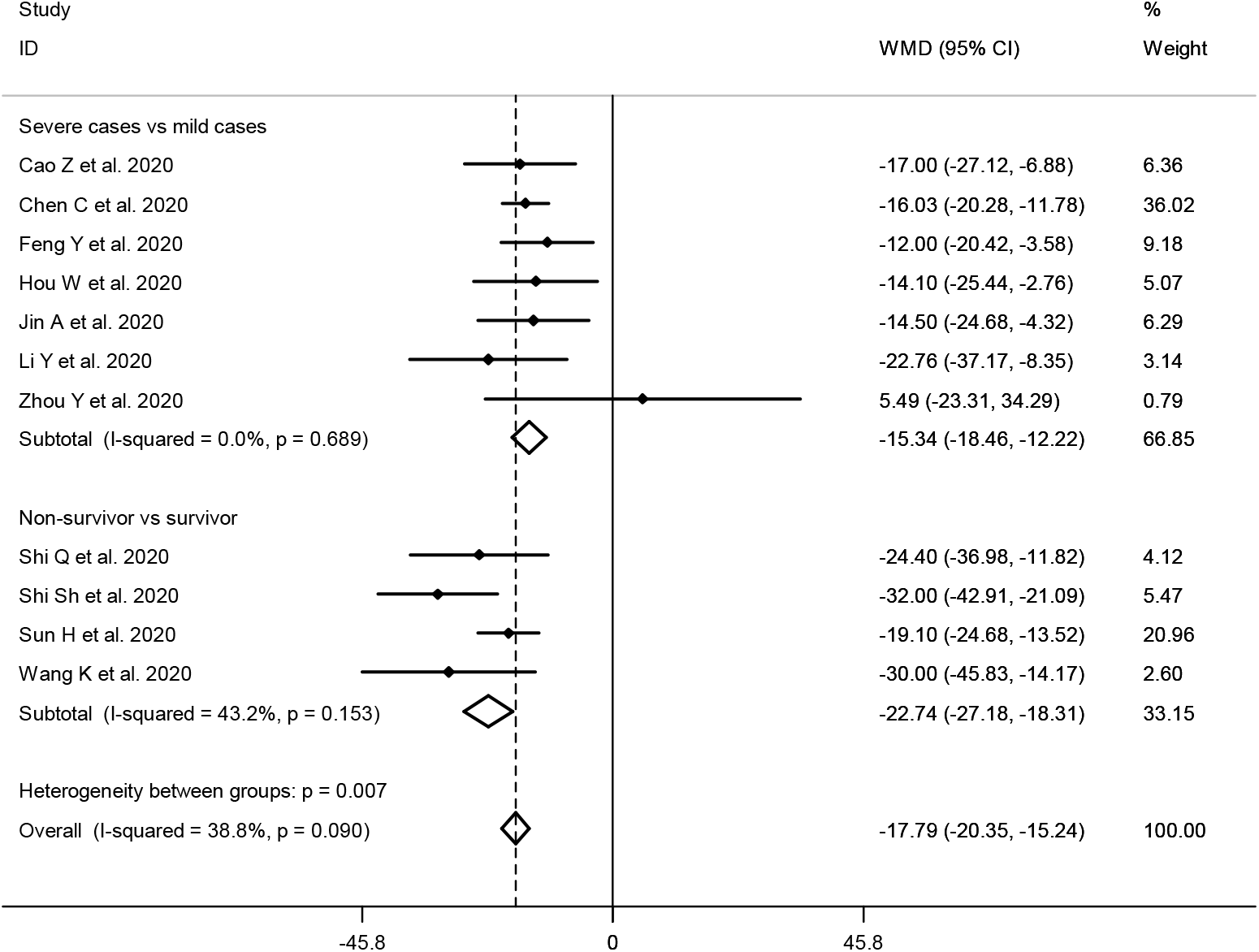
Forest plot for the association between levels of eGFR and severe outcome or death from COVID-19 infection using fixed-effects model

### Serum levels of creatinine, BUN, eGFR and mortality from COVID-19 infection

Fifteen studies ^7, 13, 16, 20, 25, 36, 41-45, 47, 48, 50, 52^ including a total of 3,080 COVID-19 infected patients (non-survivor = 775 and survivor = 2,305) reported mortality as an outcome measure. Pooled analysis showed that higher serum levels of creatinine (weighted mean difference = 18.32 μmol/L, 95% CI = 12.88 to 23.75, P<0.001, I^2^ = 64.1%, P_heterogeneity_ <0.001) (Figure 5), BUN (weighted mean difference = 3.56 mmol/L, 95% CI = 2.65 to 4.48, P<0.001, I^2^ = 72.4%, P_heterogeneity_ = 0.001) (Figure 3) and lower levels of eGFR (weighted mean difference = -22.74 mL/min/1.73 m^2^, 95% CI = -27.18 to -18.31, P<0.001, I^2^ = 43.2%, P_heterogeneity_ = 0.153) (Figure 4), were associated with a significant increase in the mortality of COVID-19 infection.

**Fig. 5.**
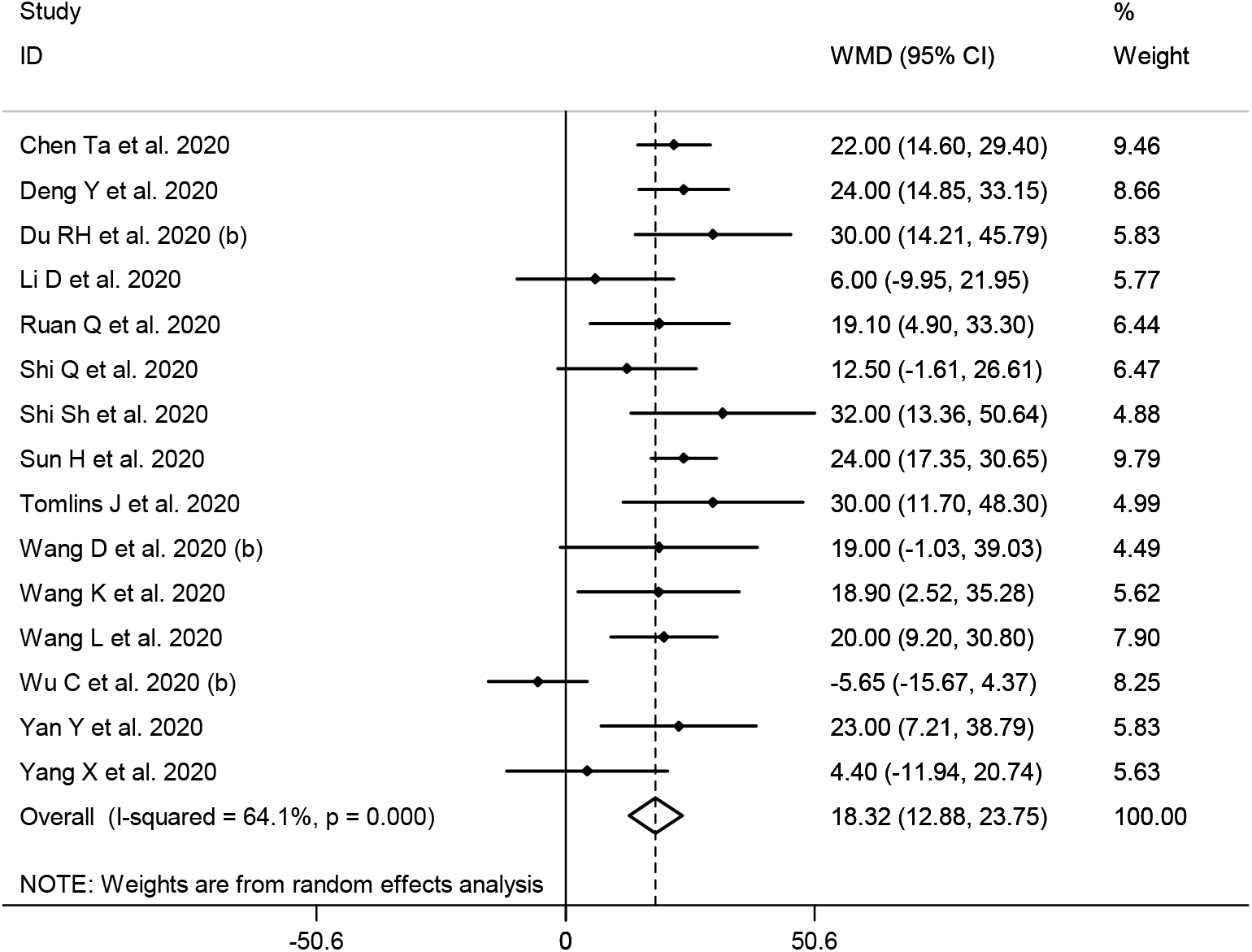
Forest plot for the association between serum levels of creatinine and mortality from COVID-19 infection using random-effects model

### Publication bias and sensitivity analysis

Based on the visual inspection of funnel plot and the results of Egger’s regression test, we found no evidence of publication bias for creatinine (P=0.924) (Supplementary Figure 1) and eGFR (P=0.612) (Supplementary Figure 2). Based on Egger's regression test, publication bias existed for BUN (P=0.024) (Supplementary Figure 3), and we used meta-trim method to re-estimate the effect size. The adjusted weighted mean difference after the trim and fill analysis was 0.73 (95% CI: 0.26 – 1.19, P = 0.002) in the random-effects model. In addition, findings from sensitivity analysis revealed that overall estimates did not depend on a single study.

### Discussion

Findings from this meta-analysis showed that kidney dysfunction is associated with severe outcome and death from COVID-19 infection. To our best knowledge, this study is the first meta-analysis to assess the association between kidney biomarkers, in particular eGFR and severity of COVID-19 infection.

The findings from our study are in agreement with previous reviews ^61-64^. Previously, kidney injury has been reported as a significant risk factor for severe outcome and death in SARS and MERS ^65, 66^.

COVID-19 may either exacerbate preexisting chronic kidney diseases and/or induce new acute kidney injuries. Earlier studies have shown that the incidence of acute kidney injury in severe COVID-19 patients and death cases ranged from 3.7% to 70.7% and 18.3% to 73.7%, respectively ^10, 25, 37, 45^.

The pathophysiology of COVID-19 associated acute kidney injury could be related to specific and nonspecific mechanisms. Specific mechanisms include direct cellular injury resulting from viral entry through the angiotensin-converting enzyme 2 receptor which is highly expressed in the kidneys ^3, 67^, an imbalanced renin–angiotensin–aldosterone system activation, leading to vasoconstriction, inflammation and fibrosis ^68, 69^, cytokine storm, mediated by increased pro-inflammatory cytokine production after COVID-19 infection ^61^ and thrombotic events ^70^. Nonspecific mechanisms include hypovolemia ^8^, right heart failure, hemodynamic changes such as central venous pressure elevation and increased intra-thoracic pressure ^71^, high levels of positive end-expiratory pressure in patients requiring mechanical ventilation ^72, 73^, nosocomial sepsis and administration of nephrotoxic drugs, especially antibiotics, antiviral therapy and/or traditional medicine ^61^.

Furthermore, increased fraction of inspired oxygen (FiO2) requirement and respiratory failure can also contribute to renal hypoperfusion and medullary hypoxia resulting from combined effects of third-space fluid loss, endothelial damage and hypotension from COVID-19-induced cytokine release ^74^.

Urine sediments of COVID-19 infected patients showed subnephrotic microhematuria and proteinuria, which are consistent with autopsy findings of kidney tissues of these patients showing proximal tubule injury with erythrocyte aggregates obstructing the peritubular capillaries and tubular necrosis with macrophage infiltration ^70^.

Until effective therapies against COVID-19 become available, the treatment of disease will be primarily based on the treatment of complications. Treatment of kidney complications should be based on optimal use of guideline-based therapies.

This study has some limitations. First, interpretation of results might be limited by the low sample size. Second, the present study did not include data such as smoking history and body mass index and, which are potential risk factors for disease severity.

### Conclusion

In this meta-analysis of 8,180 COVID-19 infected patients, acute kidney injury as assessed by serum levels of creatinine, BUN and eGFR was associated with severe outcome and death from COVID-19 infection.

## Data Availability

All data are publicly available.

## ACKNOWLEDGEMENT

None.

## Supporting Information

Supplementary table 1. Systematic literature review search terms and strategy.

Supplementary Figure 1. Funnel plot for the association between serum levels of creatinine and severity of COVID-19 infection.

Supplementary Figure 2. Funnel plot for the association between eGFR and severity of COVID-19 infection.

Supplementary Figure 3. Funnel plot for the association between BUN levels and severity of COVID-19 infection.

